# Associations of mental health with subsequent weight development during a 4-year follow-up in adolescence

**DOI:** 10.1101/2025.10.01.25337050

**Authors:** Emilia Ankkuri, Sohvi Lommi, Hanna Granroth-Wilding, Jari Lahti, Heli Viljakainen

## Abstract

**Impact:**

– This study provides up-to-date, longitudinal evidence on how mental health shapes subsequent weight development during the critical and rapid growth period of adolescence.
– Both adverse and protective dimensions of mental health play a role in shaping adolescent weight development, with evidence that some youths experiencing mental health difficulties exhibited smaller increases in BMI z-score, even when starting from a higher baseline weight status.
–It should be acknowledged that in contemporary society, adolescents with emerging mental health difficulties may not necessarily present immediate excessive weight gain but instead follow stable or even declining weight trajectories.

Although obesity and mental health problems represent two of the most pressing public health challenges, their developmental interplay remains insufficiently understood. Thus, we examined how depressive and anxiety symptoms, self-esteem, and psychological resilience are associated with weight development during adolescence.

We included 1280 11-year-old children from the Finnish Health in Teens cohort, following 814 of them for 4.3 years. Mental health was assessed with validated self-administrated scales at age 11. Psychological resilience was determined by psychological health after exposure to stressful life events. Body mass index z-score (BMIz) and waist-to-height ratio (WtHr) were calculated at ages 11 and 16.

Depressive and anxiety symptoms were associated with higher baseline BMIz but smaller increases in mean BMIz during follow-up (*p* = 0.009, *p* = 0.005, respectively). High self-esteem was associated with lower baseline BMIz, yet with steeper increases over follow-up (*p* = 0.002). Resilient adolescents showed lower baseline BMIz compared with non-resilient adolescents but exhibited greater increases during follow-up (*p* = 0.005). Similar trends were observed for WtHr.

These findings highlight that mental health is associated with physical growth in adolescence, emphasising the need to consider mental health when seeking to understand and address patterns of weight development and their underlying dynamics.

## Introduction

Obesity and mental health challenges have both reached epidemic levels, posing considerable public health concerns that increasingly affect young people.^1,2^ Roughly half of all mental disorder cases emerge during childhood and adolescence,^3^ with subclinical symptoms being even more prevalent. The Developmental Origins of Health and Disease (DOHaD) framework suggests that periods of rapid physical growth are especially sensitive to external influences and insults.^4^ Given that accelerated growth occurs during adolescence, this developmental stage may represent a critical window for the emergence and consolidation of the relationship between mental health and subsequent weight trajectories.^5^ However, the influence of mental health on physical growth and weight development during adolescence remains poorly understood.

Previous research on mental health and physical growth has primarily focused on depression and the risk of obesity, with substantial evidence supporting their co-occurence.^6–10^ Two meta-analyses summarizing longitudinal studies in adolescents concluded that particularly females with depressive symptoms have elevated risk of obesity over varying follow-up periods.^9,10^ This finding is corroborated by another systematic review.^11^ Anxiety, another negative emotional state, has received less research attention, with most existing evidence coming from cross-sectional studies.^7^ Although a few longitudinal studies suggest that anxiety in adolescence may increase the risk of consequent obesity,^11,12^ the overall literature remains scarce and inconclusive.

Poor mental health during adolescence may also manifest as weight loss or insufficient weight gain. In fact, weight loss is a diagnostic criterion for major depressive disorder and has been reported to be more common than weight gain among adolescents experiencing depression.^13,14^ One previous longitudinal study found that although affective symptoms were associated with obesity in middle age, they were also associated with a slower mean increase in BMI between ages 11 and 15 compared to those without such symptoms.^15^ However, since most earlier studies have used categorical obesity measures or examined outcomes in adulthood, they may have missed these early, adolescence-specific patterns in weight development that emerge alongside the onset of symptoms.

In addition to negative emotional states, self-esteem is a key component of mental health affecting both psychological and physical well-being.^16^ In adolescents, low self-esteem has frequently been associated with concurrent obesity.^17,18^ Although childhood low self-esteem has been associated with increased weight gain over time, and high self-esteem with a leaner body,^19,20^ these findings are not consistently supported.^21,22^ Moreover, given that much of the existing evidence is derived from earlier cohorts, their relevance may be limited in the context of today’s adolescents, who are exposed to unique and growing pressure related to body image and appearance.

Early life stress and adversity, such as parental separation, and exposure to household mental illness or substance abuse, have consistently been linked to poorer mental health and higher weight outcomes.^23^ However, not all individuals exposed to such adversity experience negative outcomes, and some can adapt and thrive despite facing substantial challenges.

Resilience, commonly defined as an individual’s ability to recover from adversity and stress,^24^ has been associated with a lower risk of mental health problems in adolescents.^25^ Cross-sectional studies have also found that higher levels of resilience are associated with a lower prevalence of obesity.^26,27^ Despite this, surprisingly little is known about how resilience relates to weight development over time. One longitudinal study in young adulthood reported that lower resilience was associated with a less favourable health profile and higher weight compared to peers with higher levels of resilience.^28^

The aim of the present study is to examine whether mental health indicators (depressive and anxiety symptoms, self-esteem and psychological resilience) associate with body mass index z-score (BMIz) or waist-to height ratio (WtHr) at the mean age of 11 years and with subsequent weight development over a four-year follow-up period. We hypothesize that adolescents who experience more mental health difficulties will exhibit BMIz and WtHr development patterns that diverge from those of their peers without such symptoms.

## Methods

### Study design and participants

This study used data from the prospective cohort study Finnish Health in Teens (www.finhit.fi). During the pilot study conducted in 2011, all adolescents born in Finland between 26 February and 6 May 2000 were recruited through mailed invitations. In total, 1599 adolescents chose to participate, yielding a participation rate of 14%. The adolescents completed a self-administered questionnaire on their anthropometric measurements, mental health, and health behaviours. The first follow-up was conducted in 2015–2016, when the adolescents self-reported their anthropometric measurements and physical activity. The participation rate at follow-up was 63%.

In the present study, we included 1280 adolescents with a mean age of 11.2 (standard deviation [SD]: 0.1) years at baseline, who met the following criteria: data were available on at least one mental health indicator at baseline (symptoms of depression, n = 1257; anxiety, n = 1267; or self-esteem, n = 1276), with no more than 50% of responses missing from the raw questionnaire items, and on anthropometric measurements at baseline (BMIz, n = 1271; WtHr, n = 1270) or follow-up (BMIz, n = 814; WtHr, n = 810). The average follow-up period length was 4.3 (±0.1) years. Of this sample, 36% of participants were lost to follow-up (n = 466). Adolescents who dropped out of the study had caregivers with significantly lower levels of education and higher BMI, had experienced a greater number of stressful life events, and were more frequently classified as either resilient or non-resilient, compared to those who remained in the study (**Supplementary Table 1**).

Written informed consent was obtained from adolescents and caregivers. The study protocol received approval from the Coordinating Ethics Committee of the Hospital District of Helsinki and Uusimaa (169/13/03/00/10).

### Measurements

#### Anthropometry at baseline and follow-up

Families were instructed to measure and report their adolescents’ height, weight, and waist circumference through a web-based survey as explained in detail elsewhere.^29^ The home-based measurements were previously validated.^30^ BMI was calculated as weight in kilograms divided by height in metres squared (kg/m^2^) and computed to an age- and sex-specific BMIz according to the International Obesity Task Force.^31^ WtHr was determined by dividing waist circumference in centimetres by height in centimetres. BMIz was used to reflect overall adiposity, whilst WtHr served as a measure for central obesity.^32^

### Indicators of mental health

All indicators of mental health were self-reported at baseline. We imputed missing scale items with a mean of existing items when the participants responded to more than 50% of the items in a scale (n = 104 to 110).

#### Depressive symptoms

Were evaluated using the 20-item Centre for Epidemiological Studies Depression Scale for Children (CES-DC).^33^ CES-DC is a self-administered questionnaire which assesses a range of depressive symptoms during the preceding week. CES-DC demonstrated a strong internal consistency in this sample, with a Cronbach’s alpha of 0.81. Previous studies have also reported a good internal consistency for CES-DC.^33^ The composite score of the scale ranged from 0 to 60, with higher scores indicating more severe symptoms.

We utilised a recommended cut-off score of ≥ 15 to categorise a clinical level of depressive symptoms.^34^

#### Anxiety symptoms

Over the preceding three months were assessed using 24 items from the Screen for Child Anxiety-Related Emotional Disorders (SCARED) child-reported version.^35^ The internal consistency of the SCARED scale was high (Cronbach’s alpha = 0.87), which is consistent with findings from previous studies.^35^ The composite score on the scale ranged from 0 to 48, with higher scores signifying a higher level of anxiety symptoms. We used a cut-off score of ≥ 22 on the SCARED scale to identify clinically significant anxiety symptoms.^36^

#### Self-esteem

Was evaluated using the 24-item Self-Perception Profile for Children (SPPC).^37^ The SPPC scale had high internal consistency (Cronbach’s alpha = 0.81), which aligns well with previous studies.^37^ The composite score on the scale ranged from 24 to 96, with higher scores reflecting a higher self-esteem. Since there are no clinical cut-offs for SPPC, belonging to the highest tertile was considered an indication of a high self-esteem.

#### Psychological resilience

Was defined as psychological well-being despite exposure to early stressful life events (SLEs), which ranged from events such as the death of a family member to the birth of a sibling.^28,38^ SLEs at baseline were assessed using the Life Events as Stressors in Childhood and Adolescence questionnaire.^39^ Participants reporting any SLEs were considered exposed, whilst those reporting no SLEs were considered unexposed.^28^ Psychological well-being encompassed not having clinically significant symptoms of depression or anxiety (i.e., psychological distress) evaluated using CES-DC and SCARED, respectively, or high self-esteem score, as measured by the SPPC. Psychological resilience was categorised into four phenotypes: resilient (no clinically relevant depressive or anxiety symptoms, or high self-esteem, after exposure to SLEs), non-resilient (clinically significant symptoms of depression or anxiety, and low self-esteem, after exposure to SLEs), psychological well-being without adversity (unexposed to SLEs, no clinically significant symptoms of depression or anxiety, or high self-esteem), and psychological distress without adversity (unexposed to SLEs, clinically significant symptoms of depression or anxiety, and low self-esteem).

### Covariates

Personal information regarding age and sex was obtained from the Population Information System at the Population Register Centre. Adolescents reported their usual leisure time physical activity (in hours/week) at baseline and follow-up through a questionnaire. Puberty status was self-evaluated by adolescents using the five-point Tanner scale^40^ and categorised into prepubertal, pubertal, or post-pubertal. Based on a self-administered food propensity questionnaire at baseline, adolescents were clustered as having a dietary pattern defined as healthy, unhealthy, or avoiding fruit and vegetable.^41^ At baseline, caregiver was asked to report their highest level of educational qualification, which was categorised into lower (comprehensive school), middle (vocational school or equivalent), and higher level (university degree or equivalent). Caregivers’ BMIs was calculated based on their self-reported height and weight at baseline and at follow-up.

### Statistical analyses

Statistical analyses were calculated using R studio, version 2022.12.0+353. We considered *p* < 0.05 as statistically significant. BMIz and WtHr exhibited a satisfactory normal distribution (skewness < 1.8).

Using a linear regression model, we examined the cross-sectional associations of mental health indicators with BMIz and WtHr at baseline. Model 1 was adjusted for sex and age, while Model 2 was additionally adjusted for physical activity, dietary pattern, puberty status, caregivers’ BMI and caregivers’ education level.

To study the longitudinal associations between baseline mental health and changes in BMIz and WtHr during the follow-up period, we employed linear mixed models (LMMs). For these analyses, we centred continuous depressive symptoms, anxiety symptoms, self-esteem, and age. We fitted random intercepts to allow the average BMIz and WtHr to vary between individuals. Mental health indicators were included as fixed effects, and their interactions with age were considered to examine how the association of mental health indicators with BMIz and WtHr changes with age. To illustrate the results of the LMMs, participants were categorized based on their scores on each continuous mental health indicator into three groups: low (< mean - 1SD), average, and high (> mean + 1SD). Pairwise comparisons between groups were then calculated using the *emmeans* package in R, with all *p*-values presented without adjustment for multiple comparisons.

## Results

### Participants characteristics

Our sample consisted of 1280 participants (51% girls) with an average (SD) age of 11.2 (0.1) at baseline. Participant characteristics are presented in **Table 1**. In the entire sample, 17% presented with overweight or obesity and 15% with central obesity at baseline. In the subsample with follow-up data available, 15% lived with overweight or obesity and 9% with central obesity at follow-up. The mean BMIz increased by 0.09 units (*p* < 0.001), while mean WtHr decreased by 0.01 units (*p* < 0.001) over the follow-up period.

**Table 1.**
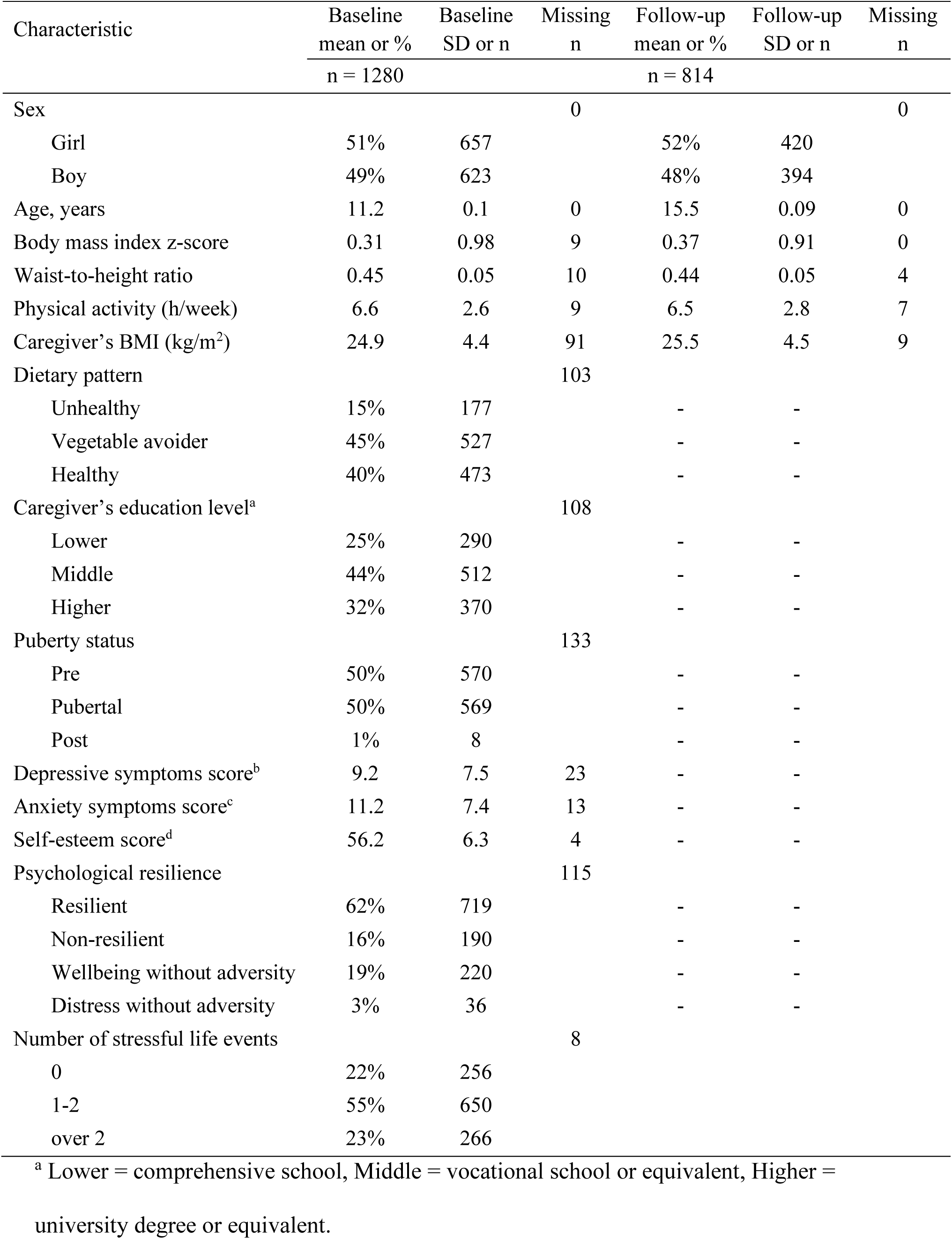
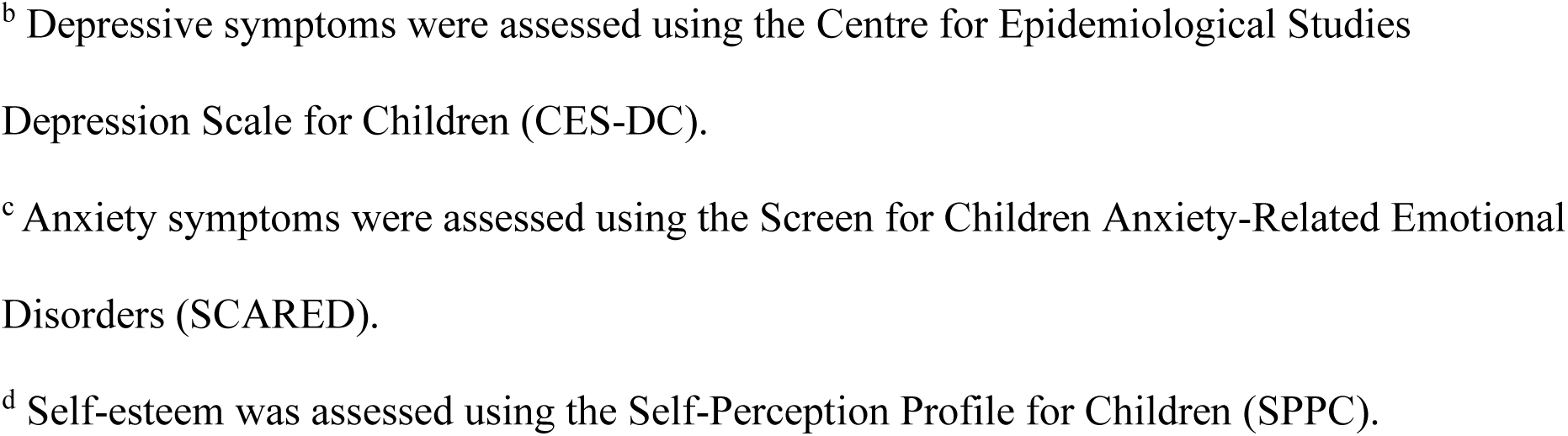
Characteristics of the sample at baseline and follow-up.

| Characteristic | Baseline<br>mean or %<br>n = 1280 | Baseline<br>SD or n | Missing<br>n | Follow-up<br>mean or %<br>n = 814 | Follow-up<br>SD or n | Missing<br>n |
| --- | --- | --- | --- | --- | --- | --- |
| Sex |  |  | 0 |  |  | 0 |
| Girl | 51% | 657 |  | 52% | 420 |  |
| Boy | 49% | 623 |  | 48% | 394 |  |
| Age, years | 11.2 | 0.1 | 0 | 15.5 | 0.09 | 0 |
| Body mass index z-score | 0.31 | 0.98 | 9 | 0.37 | 0.91 | 0 |
| Waist-to-height ratio | 0.45 | 0.05 | 10 | 0.44 | 0.05 | 4 |
| Physical activity (h/week) | 6.6 | 2.6 | 9 | 6.5 | 2.8 | 7 |
| Caregiver's BMI (kg/m<sup>2</sup>) | 24.9 | 4.4 | 91 | 25.5 | 4.5 | 9 |
| Dietary pattern |  |  | 103 |  |  |  |
| Unhealthy | 15% | 177 |  | - | - |  |
| Vegetable avoider | 45% | 527 |  | - | - |  |
| Healthy | 40% | 473 |  | - | - |  |
| Caregiver's education level<sup>a</sup> |  |  | 108 |  |  |  |
| Lower | 25% | 290 |  | - | - |  |
| Middle | 44% | 512 |  | - | - |  |
| Higher | 32% | 370 |  | - | - |  |
| Puberty status |  |  | 133 |  |  |  |
| Pre | 50% | 570 |  | - | - |  |
| Pubertal | 50% | 569 |  | - | - |  |
| Post | 1% | 8 |  | - | - |  |
| Depressive symptoms score<sup>b</sup> | 9.2 | 7.5 | 23 | - | - |  |
| Anxiety symptoms score<sup>c</sup> | 11.2 | 7.4 | 13 | - | - |  |
| Self-esteem score<sup>d</sup> | 56.2 | 6.3 | 4 | - | - |  |
| Psychological resilience |  |  | 115 |  |  |  |
| Resilient | 62% | 719 |  | - | - |  |
| Non-resilient | 16% | 190 |  | - | - |  |
| Wellbeing without adversity | 19% | 220 |  | - | - |  |
| Distress without adversity | 3% | 36 |  | - | - |  |
| Number of stressful life events |  |  | 8 |  |  |  |
| 0 | 22% | 256 |  |  |  |  |
| 1-2 | 55% | 650 |  |  |  |  |
| over 2 | 23% | 266 |  |  |  |  |

<sup>a</sup> Lower = comprehensive school, Middle = vocational school or equivalent, Higher = university degree or equivalent.
<sup>b</sup> Depressive symptoms were assessed using the Centre for Epidemiological Studies Depression Scale for Children (CES-DC).
<sup>c</sup> Anxiety symptoms were assessed using the Screen for Children Anxiety-Related Emotional Disorders (SCARED).
<sup>d</sup> Self-esteem was assessed using the Self-Perception Profile for Children (SPPC).

Clinically significant symptoms of depression and anxiety based on recommended cut-off scores at baseline were observed in 18% and 9% of participants, respectively. Moreover, 62% were defined as resilient and 16% as non-resilient, whilst 3% had a low psychological well-being without adversity, and 19% were defined as having high well-being without adversity. Of the sample, 78% reported at least one stressful life event (SLEs) during their lifetime. The most reported SLEs were the birth of a sibling (45%) and family relocation (30%) (**Supplementary Table 2**).

### Associations between mental health indicators and anthropometry at baseline

The cross-sectional associations between mental health indicators and BMIz baseline are presented in **Table 2**. After adjusting for sex and age (Model 1), higher levels of both depressive and anxiety symptoms were associated with higher BMIz (b = 0.008, *p* = 0.03 and b = 0.007, *p* = 0.05, respectively). After adjusting for physical activity, dietary pattern, puberty status, caregivers’ BMI and caregivers’ education level in addition to the Model 1 covariates (Model 2), these associations were no longer statistically significant (*p*-value > 0.1). Higher self-esteem was associated with lower BMIz (b = −0.028, *p* < 0.001) in Model 1, and these associations remained significant in the Model 2 (b = −0.02, *p* < 0.001). Compared with their non-resilient peers, adolescents classified as psychologically resilient had lower BMIz (b = −0.163, *p* = 0.040) in Model 1. In Model 2, the association with BMIz was no longer statistically significant (*p*-value > 0.1). Similar patterns of associations were observed for WtHr (**Supplementary Table 3**).

**Table 2.** Cross-sectional linear associations of mental health indicators with body mass index z-score (BMIz) at age 11 (baseline), indicated with unstandardized b coefficients with 95% confidence intervals (CI).

| Mental health indicator | Model 1: b [95% CI] | p-value | df | Model 2: b [95% CI] | p-value | df |
| --- | --- | --- | --- | --- | --- | --- |
| Depressive symptoms | 0.008 [0.001, 0.015] | **0.027** | 1244 | 0.006 [-0.002, 0.014] | 0.13 | 956 |
| Anxiety symptoms | 0.007 [0, 0.015] | **0.05** | 1254 | 0.003 [-0.005, 0.011] | 0.43 | 954 |
| Self-esteem | -0.028 [-0.036, -0.019] | **<0.001** | 1263 | -0.020 [-0.029, -0.010] | **<0.001** | **954** |
| Psychological resilience<sup>a</sup> |  |  | 1153 |  |  | 929 |
| Resilient | -0.163 [-0.318, -0.007] | **0.040** |  | -0.116 [-0.279, 0.043] | 0.17 |  |
| Wellbeing without adversity | -0.116 [-0.305, 0.073] | 0.23 |  | -0.067 [-0.262, 0.130] | 0.51 |  |
| Distress without adversity | -0.216 [-0.562, 0.129] | 0.22 |  | -0.281 [-0.644, 0.083] | 0.13 |  |

Model 1 adjusted for sex and age. Model 2 adjusted for sex, age, puberty status, physical activity, dietary pattern, caregivers' education level, and caregiver's BMI.
<sup>a</sup> Reference = non-resilient group. Coefficients for other groups represent the differences relative to this group.

### Associations between mental health indicators and change in anthropometry Depressive and anxiety symptoms

In the LMMs investigating depressive and anxiety symptoms, age showed a significant positive main effect, indicating that BMIz increased with age during the follow-up (**Table 3**). However, we found significant negative interactions between age and symptoms of depression and anxiety on BMIz measurements (*p* for interactions between symptoms*age < 0.05; **Table 3**, **Figure 1: a1, a2**). After adjusting for covariates in Model 2, the interaction between depressive symptoms and age remained statistically significant, though attenuated (b = −0.002, *p* = 0.03), whereas the interaction with anxiety symptoms was no longer significant (*p* > 0.05).

**Figure 1.**
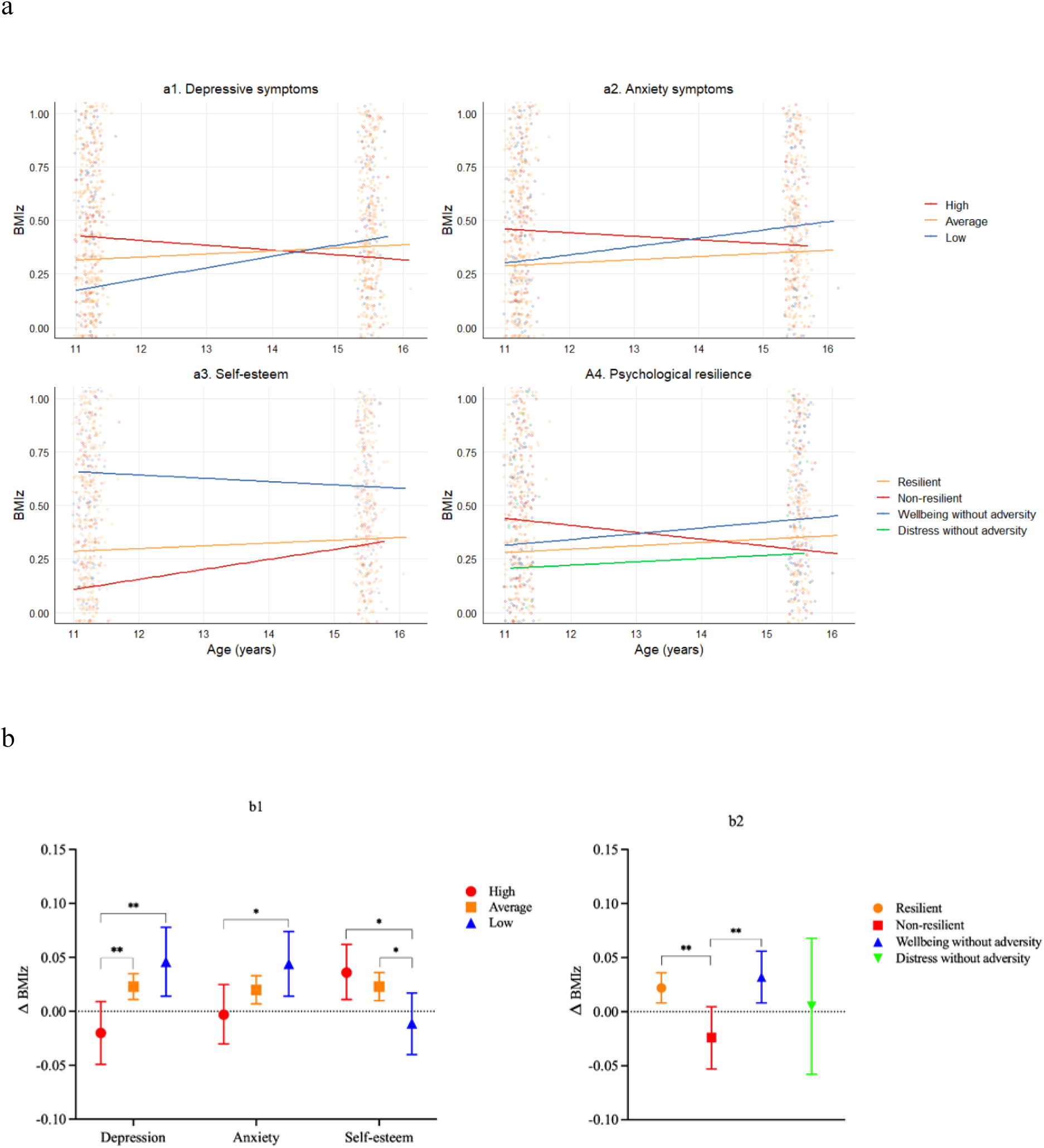
Panels a1–a4 illustrate the interaction between age x (A1) depressive symptoms, (A2) anxiety symptoms, (A3) self-esteem, or (A4) psychological resilience predicting changes in BMIz, corresponding to the terms reported in Table 3. Panels B1 and B2 present yearly changes in BMIz with 95% confidence intervals, alongside pairwise group comparisons: low, average and high levels of depressive and anxiety symptoms, self-esteem (B1) and different resilience groups (B2). Pairwise comparisons between the low, average, and high groups were conducted using the emmeans package in R (see Table 4). *p<0.05, **p<0.01. For depressive symptoms, anxiety symptoms, and self-esteem, participants were grouped into low (mean - 1 SD), average, and high (mean +1 SD). Individual raw data points are overlaid in panels A1-A4 to illustrate distribution of observations. The y-axis range is restricted to 0.00-1.00 in panel A and −0.10-0.015 in panel B to better illustrate the modest differences over time.

**Table 3.**
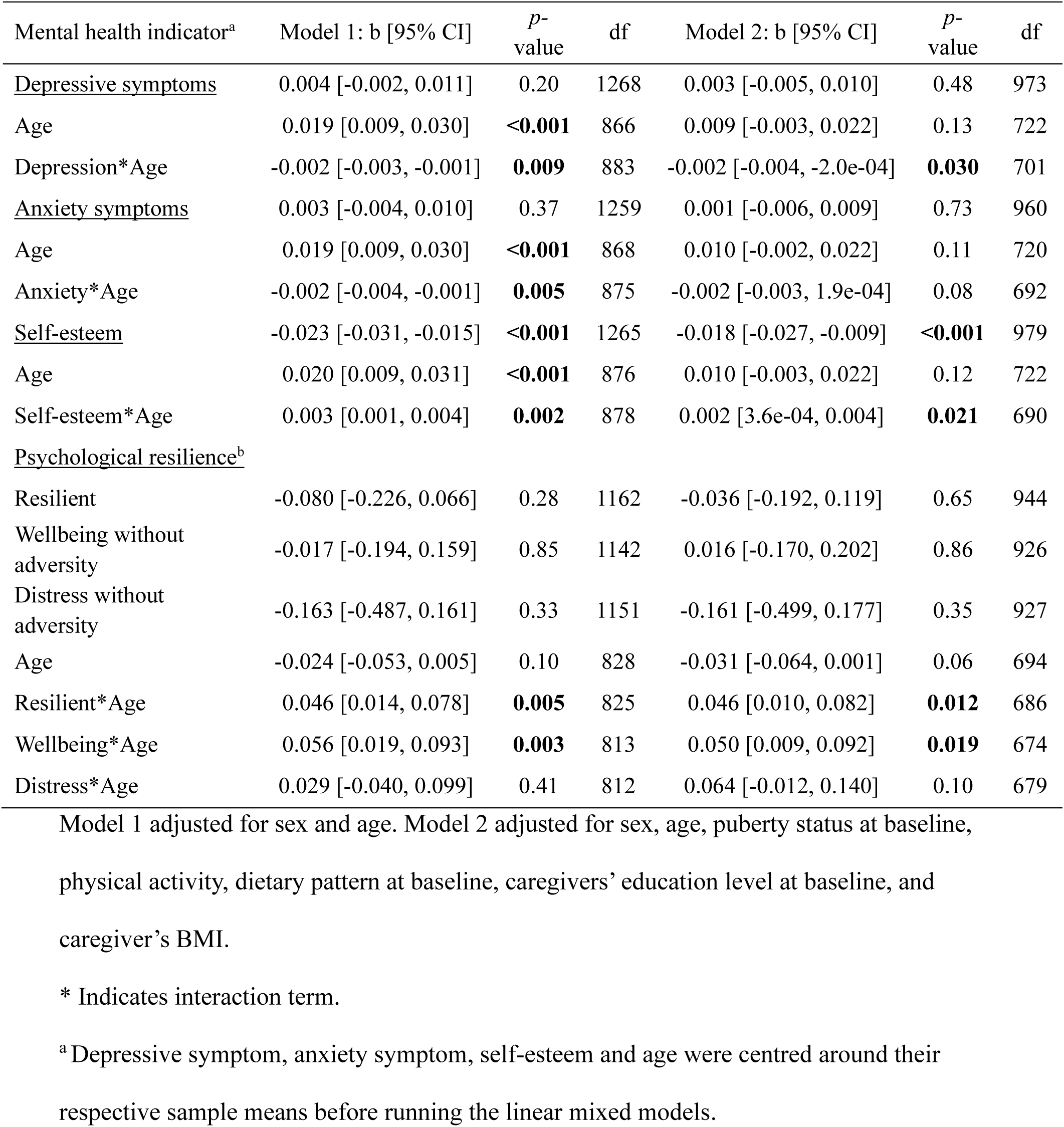
Associations between mental health indicators at age 11 and changes in body mass index z-scores (BMIz) over a 4.3-year follow-up period, indicated with unstandardized fixed effect regression coefficients (b) with standard errors and 95% confidence intervals (CI) from linear mixed models.

**Table 4.** Results from pairwise comparisons of groups according to levels of depression and anxiety symptoms, self-esteem and psychological resilience on change between baseline and follow-up in BMIz (Δ BMIz).

| Mental health indicator | Comparison | Model 1:<br>ΔBMIz | *p*-value | Model 2:<br>ΔBMIz | *p*-value |
| --- | --- | --- | --- | --- | --- |
| Depressive symptoms |  |  |  |  |  |
|  | Low vs. Average | 0.023 | 0.19 | 0.032 | 0.10 |
|  | High vs. Average | -0.043 | **0.007** | -0.037 | **0.043** |
|  | High vs. Low | -0.066 | **0.003** | -0.069 | **0.006** |
| Anxiety symptoms |  |  |  |  |  |
|  | Low vs. Average | 0.024 | 0.16 | 0.014 | 0.45 |
|  | High vs. Average | -0.023 | 0.14 | -0.020 | 0.24 |
|  | High vs. Low | -0.046 | **0.026** | -0.035 | 0.14 |
| Self-esteem |  |  |  |  |  |
|  | Low vs. Average | -0.035 | **0.029** | -0.050 | **0.008** |
|  | High vs. Average | 0.013 | 0.36 | 0.002 | 0.90 |
|  | High vs. Low | 0.048 | **0.014** | 0.052 | **0.021** |
| Psychological resilience |  |  |  |  |  |
|  | Resilient vs. Non-resilient | 0.046 | **0.005** | 0.046 | **0.012** |
|  | Resilient vs. Wellbeing | -0.011 | 0.46 | -0.004 | 0.78 |
|  | Resilient vs. Distress | 0.016 | 0.62 | -0.017 | 0.63 |
|  | Non-resilient vs. Wellbeing | -0.056 | **0.003** | -0.050 | **0.019** |
|  | Non-resilient vs. Distress | -0.029 | 0.41 | -0.064 | 0.10 |
|  | Wellbeing vs. Distress | 0.027 | 0.44 | -0.013 | 0.73 |

$\Delta$ = Difference in b coefficient.
Low group = mean - 1 SD, High group = mean + 1 SD.
Model 1 adjusted for sex and age. Model 2 adjusted for sex, age, puberty status at baseline, physical activity, dietary pattern at baseline, caregivers' education level at baseline, and caregiver's BMI.

Pairwise comparisons of estimated marginal trends revealed differences in yearly BMIz change across groups according to low, average and high levels of symptoms (**Table 4**, **Figure 1: b1**). In Model 1, adolescents with high depressive symptoms exhibited a slower increase in BMIz compared to those with low (difference = −0.066, *p* = 0.003) and average (difference = −0.043, *p* = 0.007) symptom levels. This pattern was consistent in Model 2, suggesting a more gradual change in BMIz among adolescents with higher depressive symptoms relative to other groups (difference = −0.069, *p* = 0.006 *for high vs. low*; difference = −0.037, *p* = 0.043 *for high vs. average*). Similar, though less pronounced differences were observed between the high and low anxiety symptom groups in Model 1 (difference = −0.046, *p* = 0.026), however, this was no longer significant after full adjustment in Model 2 (*p* > 0.1). Comparable patterns were observed for WtHr in relation to depressive and anxiety symptoms (**Supplementary Table 4-5, Supplementary Figure 1**). The steepest decrease in WtHr over the follow-up period was observed in those with high levels of symptoms.

### Self-esteem

For self-esteem, age showed a positive overall effect on BMIz (**Table 3**). In contrast to depressive and anxiety symptoms, the interaction between age and self-esteem on BMIz was positive (*p* for interaction self-esteem*age < 0.05; **Table 3**, **Figure 1: a3**). After adjusting for covariates in Model 2, the interaction between self-esteem and age remained statistically significant, thought attenuated slightly (b= 0.0002, *p* = 0.021). Again, we illustrated the interaction by examining changes in mean BMIz across groups with high, average, and low levels of self-esteem (**Table 4**, **Figure 1: b1**). In the pairwise comparisons, the change in BMIz differed significantly between the high and low groups and between the average and low groups: steeper increases in BMIz over time occurred in high and average groups compared to the low self-esteem group (difference = 0.048, *p* = 0.014 *for high vs. low*; difference = 0.035, *p* = 0.029 *for average vs low*). These associations remained significant in Model 2. Comparable patterns were observed for WtHr (**Supplementary Table 4-5, Supplementary Figure 1**).

### Psychological resilience

A significant interaction between psychological resilience groups and age was observed (*p* for interactions between resilient and wellbeing without adversity groups*age < 0.05; **Table 3**, **Figure 1: a4**). In contrast to the non-resilient group, the resilient group showed an increase in BMIz over time (b = 0.046, *p* = 0.005). Pairwise comparisons of estimated marginal trends confirmed that the resilient group experienced a steeper increase in BMIz compared to the non-resilient group (difference = 0.046, *p* = 0.005; **Table 4**, **Figure 1: b2**). Furthermore, the non-resilient group had a more attenuated change in BMIz compared to the wellbeing without adversity group (difference = −0.056, *p* = 0.003). Further adjustments in Model 2 had minimal impact on these results. Regarding WtHr, all groups demonstrated decrease in WtHr over time (**Supplementary Table 4-5, Supplementary Figure 1**). However, the non-resilient group showed a more pronounced decline compared to the resilient group.

## Discussion

We demonstrated that both risk and protective aspects of mental health were associated with subsequent changes in BMIz and WtHr in a contemporary adolescent cohort. A modest increase in mean BMIz was observed across the cohort during the four-year follow-up, however the pattern of this change differed depending on the severity of mental health symptoms. While depressive and anxiety symptoms were linked to higher BMIz at age 11, the negative interaction observed in the longitudinal models indicated that these associations weakened over the course of the four-year follow-up. In contrast, adolescents with higher self-esteem and psychological resilience showed lower baseline BMIz yet experienced the most pronounced increases in BMIz over the subsequent four years. Similar trends were noted for waist-to-height ratio, which followed parallel patterns to those of BMIz over time.

We found that negative emotional states, particularly depressive symptoms, were associated with attenuated BMIz development. To our knowledge, this is the first study to demonstrate such BMIz development among adolescents born in the 2000s. While previous studies have predominantly reported ongoing excessive weight gain among adolescents with psychological distress,^11,42^ our findings point to a potentially distinct developmental trajectory, particularly for those with highest levels of depressive symptoms at early adolescence. Our findings are in line with a few earlier studies reporting that depressive symptoms may be associated with attenuated weight gain between the ages 11 and 15 compared to adolescents with fewer symptoms.^13,15^ Our findings indicate that depressive symptoms during adolescence may not drive continued excessive weight gain from ages 11 to 16. Instead, adolescents with lower levels of symptoms tended to catch up and even surpass BMIz of those with higher levels of symptoms.

In the present study, self-esteem and psychological resilience showed inverse associations with BMIz and WtHr during adolescence, in contrast to negative emotional states. Relative to their peers, adolescents with higher self-esteem and those belonging to the resilient group had lower BMIz at age 11 and experienced a more rapid increase in BMIz over the four-year follow-up period. These findings are consistent with previous literature highlighting self-esteem as a key protective factor, often showing inverse associations compared to depressive symptoms.^17^ In line with earlier research, lower self-esteem at age 11 was associated with higher concurrent BMIz. While the gap in BMIz between low and high self-esteem groups narrowed during adolescence, the groups did not fully converge, and the differences established in early adolescence remained evident throughout the follow-up period. Notably, and in contrast to some prior findings,^20^ we did not observe that low self-esteem contributed to a progressive or excessive increase in BMIz during adolescence.

Our findings regarding psychological resilience align with limited previous studies,^26–28^ showing that those adolescents classified as resilient tend to follow BMIz development patterns similar to those with high self-esteem. Moreover, our results highlight the possibility that the relationship between resilience as well as self-esteem and BMIz development may be developmentally sensitive. During periods marked by expected physiological growth and weight gain, such as between ages of 11 and 16, resilience may reflect adaptive functioning that supports normative physical development. Conversely, lower resilience might limit an adolescent’s ability to cope with physical and emotional changes, potentially contributing to deviations from healthy weight gain trajectories.

The observed BMIz differences in our study were relatively small and generally remained within the normal, healthy weight range. Adolescents with higher levels of depressive symptoms gained approximately 20.6 kg over four years, whereas those with lower symptoms levels gained around 22.6 kg. Although a 2-kg difference may appear minor, it is important to consider the short duration of follow-up. If such trajectories were to persist over decades, these differences could accumulate into clinically meaningful weight disparities. Furthermore, if these weight development patterns progress towards rapid weight gain in later adolescence, as suggested by previous studies,^15^ the trend we observed may serve as an early indicator for the timing of targeted intervention.

The link between mental health and weight changes during adolescence is complex and likely influenced by multiple social, psychological and physiological factors. Social aspects such as lack of relationships, social isolation, and reduced time spent with peers may lead to changes in eating habits, which can affect weight development. Puberty is a critical developmental period marked by normal, gradual increases in BMI as part of typical growth.^43^ Adolescents with good mental health may better accept these bodily changes, reducing body dissatisfaction and preventing unhealthy behaviors.^44^ Conversely, poor mental health has been linked to behaviours such as altered appetite, disturbed sleep, excessive screen time, and physical inactivity, all of which can promote unhealthy weight changes.^11^ Although we adjusted for lifestyle factors, residual confounding may still exist in our findings. Due to the observational study design, we were unable to examine the underlying mechanisms in detail. Further research is needed to clarify these pathways.

The strengths of the present study include its prospective design with a four-year follow-up period during critical period in adolescence. In addition, the use of a geographically diverse Finnish cohort born in the 2000s enhances the contemporary relevance of the findings. The questionnaire was extensive, allowing for the exploration of both risk and protective aspects of mental health, assessed using validated and reliable scales. Given our rich dataset, we could adjust for some potential confounding factors that could partly explain some of the observed associations. Furthermore, we used age- and sex-specific BMIz as well as WtHr to reflect adiposity. These continuous outcome measures allowed us to more precisely and comprehensively examine associations than would be possible using only categorical outcomes. In addition, employing multilevel modelling as a statistical method provides advantages over a simple linear regression model, such as accounting for individual differences in physical growth.

Simultaneously, our results should be considered in the context of certain limitations. Firstly, the response rate was relatively low (14%), which may introduce selection bias and limit the generalizability of the results. It is possible that families who chose to participate differed systematically from those who did not. As highlighted by the attrition analysis, the subsample with available follow-up data originated from parents with higher educational backgrounds than those who were lost from the follow-up.^29^ The overall high educational level amongst caregivers combined with the low rate of overweight/obesity amongst adolescents implies a possible selection bias, particularly given that obesity and mental health issues are typically more prevalent in disadvantaged socioeconomic groups.^45^ However, the youth in our study showed similar levels of depression and anxiety symptoms and rates of overweight when compared with those reported in the 2017 Finnish School Health Promotion Study at the end of comprehensive school.^46^ Since most participants remained within healthy BMIz ranges throughout the study period, this may have attenuated the associations between mental health and anthropometric measurements. Secondly, we relied on self-reported anthropometric measurements, potentially leading to limitations in reliability and accuracy.^47^ However, we previously reported the reliability of home-assessed height and weight and considered them fit for epidemiological studies.^30^ Thirdly, we assessed mental health only at the beginning of the study. To gain a nuanced picture of the associations between mental health and physical growth over time, we would need regular assessments of mental health as well as anthropometric measurements. Our study only measured anthropometric at two time points over the four-year follow-up. This may have limited our ability to capture the dynamic of weight development, which could be apparent with more frequent measurements. It is worth noting that some of the observed changes may reflect baseline differences in anthropometry. While the random effects model accounts for individual average responses, it does not fully adjust for baseline levels. Moreover, we have measured depressive and anxiety symptoms which are not equivalent to psychiatric diagnoses.

To conclude, this study provides up-to-date insights into how mental health influences weight development during a critical period of physical and psychological growth. In this contemporary adolescent cohort, mental health indicators assessed at age 11 were associated with subsequent weight development. Adolescents experiencing psychological distress, particularly depressive symptoms, showed less pronounced weight gain compared to those with lower levels of such symptoms, while the opposite pattern was observed among those with high self-esteem and psychological resilience. Although modest in magnitude, these effects may have important implications during this critical developmental period, shaping both physical and psychological trajectories across the life course.

## Data availability

The datasets generated and analysed during the current study are available from the corresponding author on reasonable request.

## Acknowledgements

We sincerely thank all participants of the Fin-HIT study, along with the school staff, fieldworkers, and research coordinators who assisted with data collection.

## Author contributions

All authors have made a substantial contribution to the work being presented. JL and HV conceptualised the study. EA performed the literature search and data analysis with support from JL and HV. HGW assisted in interpreting the results. EA prepared the initial draft of the manuscript. All authors contributed to interpreting the results and editing and revising the manuscript and approved the final version for submission.

## Consent statement

Written informed consent was obtained from Finnish Health in Teens participating adolescents and their caregivers.

