## Supporting Information for "Associations of mental health with subsequent weight development during a 4-year follow-up in adolescence"

**Title:**

**Departments:**

[1]Folkhälsan Research Centre, Helsinki, Finland

[2]Faculty of Medicine, University of Helsinki, Helsinki, Finland

[3]Department of Psychology and Logopedics, Faculty of Medicine, University of Helsinki, Helsinki, Finland

These authors contributed equally: Jari Lahti, Heli Viljakainen

***Corresponding author:**

Correspondence to Heli Viljakainen

Folkhälsan Research Centre

Topeliuksenkatu 20, 00250 Helsinki

+358 40 5916998

**Keywords (3-6):**

depression, anxiety, self-esteem, resilience, obesity, paediatrics

**Supplementary Table 1.** Comparison of baseline characteristics between participants who completed the follow-up (64%) and those who were lost to follow-up (36%).

| Characteristic | Follow-up mean (SD) or % | Missing n | Drop out mean (SD) or % | Missing n | p-value[a] |
|---|---|---|---|---|---|
| | n = 814 | | n = 466 | | |
| Sex | | 0 | | 0 | 0.84 |
| Girl | 52% | | 51% | | |
| Boy | 48% | | 49% | | |
| Age, years | 11.23 (0.11) | 0 | 11.24 (0.11) | 0 | **0.036** |
| Body mass index z-score | 0.28 (0.95) | 7 | 0.38 (1.02) | 2 | 0.09 |
| Waist-to-height ratio | 0.45 (0.05) | 7 | 0.45 (0.05) | 4 | 0.25 |
| Physical activity (h/week) | 6.6 (2.7) | 6 | 6.8 (2.6) | 3 | 0.13 |
| Caregiver's BMI (kg/m$^2$) | 24.74 (4.27) | 50 | 25.27 (4.61) | 42 | **0.049** |
| Dietary pattern | | 57 | | 47 | 0.59 |
| Unhealthy | 14% | | 16% | | |
| Vegetable avoider | 45% | | 45% | | |
| Healthy | 41% | | 39% | | |
| Caregiver's education level[b] | | 70 | | 43 | **0.010** |
| Lower | 22% | | 30% | | |
| Middle | 45% | | 42% | | |
| Higher | 33% | | 29% | | |
| Puberty status | | 65 | | 68 | 0.999 |
| Pre | 50% | | 50% | | |
| Pubertal | 50% | | 49% | | |
| Post | 0% | | 1% | | |
| Depressive symptoms score[c] | 8.9 (7.1) | 8 | 9.6 (8.1) | 15 | 0.19 |
| Anxiety symptoms score[d] | 11.2 (7.2) | 6 | 11.4 (7.7) | 7 | 0.69 |
| Self-esteem score[e] | 56.4 (6.2) | 1 | 55.9 (6.4) | 3 | 0.15 |
| Psychological resilience | | 54 | | 61 | **0.008** |
| Resilient | 61% | | 64% | | |
| Non-resilient | 15% | | 19% | | |
| Wellbeing without adversity | 22% | | 14% | | |
| Distress without adversity | 3% | | 3% | | |
| Number of stressful life events | | 51 | | 57 | **<0.001** |
| 0 | 25% | | 17% | | |
| 1–2 | 55% | | 56% | | |
| over 2 | 20% | | 28% | | |

[a] Results from Welch's t-tests (continuous variables) or Pearson's chi-square test (categorical variables).

[b] Lower = comprehensive school, Middle = vocational school or equivalent, Higher = university degree or equivalent.

[c] Depressive symptoms were assessed using the Center for Epidemiological Studies Depression Scale for Children (CES-DC).

[d] Anxiety symptoms were assessed using the Screen for Children Anxiety-Related Emotional Disorders (SCARED).

[e] Self-esteem was assessed using the Self-Perception Profile for Children (SPPC).

**Supplementary Table 2**. Prevalence of stressful life events measured by the Life Events as Stressors in Childhood and Adolescence questionnaire among participants (n = 1,280).

| Life event | Has not happened (%) | Has happened (%) | Missing (n) |
|---|---|---|---|
| Family moved to another municipality | 70% | 30% | 41 |
| Sibling was born | 55% | 45% | 61 |
| Family member was seriously ill/injured | 80% | 20% | 52 |
| Parental separation or divorce | 80% | 20% | 41 |
| Parents argued with each other more than previously | 80% | 20% | 50 |
| Mother/father became unemployed | 89% | 11% | 46 |
| Family member died | 92% | 8% | 51 |
| Parent was accused, arrested or convicted of a crime | 99% | 1% | 48 |
| Family had financial difficulties | 89% | 11% | 59 |

**Supplementary Table 3.** Cross-sectional linear associations of mental health indicators with waist-to-height ratio (WtHr) at age 11 (baseline), indicated with unstandardized b coefficients with 95% confidence intervals (CI).

| Mental health indicator | Model 1: b [95% CI] | *p*-value | df | Model 2: b [95% CI] | *p*-value | df |
|---|---|---|---|---|---|---|
| Depressive symptoms | 0.001 [3.3e-04, 0.001] | **<0.001** | 1242 | 0.001 [2.2e-04, 0.001] | **0.002** | 955 |
| Anxiety symptoms | 0.001 [3.5e-04, 0.001] | **<0.001** | 1252 | 0.001 [1.9e-04, 0.001] | **0.004** | 953 |
| Self-esteem | -0.002 [-0.002, -0.001] | **<0.001** | 1261 | -0.001 [-0.002, -0.001] | **<0.001** | 953 |
| Psychological resilience[a] | | | 1152 | | | 927 |
| Resilient | -0.012 [-0.020, -0.005] | **0.002** | | -0.010 [-0.018, -0.002] | **0.017** | |
| Wellbeing without adversity | -0.014 [-0.023, -0.004] | **0.004** | | -0.012 [-0.022, -0.002] | **0.018** | |
| Distress without adversity | -0.011 [-0.028, 0.006] | 0.20 | | -0.018 [-0.036, 0.001] | 0.06 | |

Model 1 adjusted for sex and age. Model 2 adjusted for sex, age, puberty status, physical activity, dietary pattern, caregivers' education level, and caregiver's BMI.

[a] Reference = non-resilient group. Coefficients for other groups represent the differences relative to this group.

**Supplementary Table 4.** Associations between mental health indicators at age 11 and changes in waist-to-height ratio (WtHr) over a 4.3-year follow-up period, indicated with unstandardized fixed effect regression coefficients (b) with standard errors and 95% confidence intervals (CI) from linear mixed models.

| Mental health indicator[a] | Model 1: b [95% CI] | *p*-value | df | Model 2: b [95% CI] | *p*-value | df |
|---|---|---|---|---|---|---|
| Depressive symptoms | 0.0004 [0.0001, 0.0008] | **0.011** | 1248 | 0.0003 [-3.6-05, 0.0007] | 0.08 | 954 |
| Age | -0.0024 [-0.0030, -0.0018] | **<0.001** | 870 | -0.0027 [-0.0034, -0.0021] | **<0.001** | 714 |
| Depression*Age | -0.0001 [-0.0002, -0.0001] | **0.002** | 891 | -0.0002 [-0.0002, -0.0001] | **0.002** | 701 |
| Anxiety symptoms | 0.0005 [0.0002, 0.0008] | **0.004** | 1236 | 0.0004 [1.8-05, 0.0008] | **0.041** | 939 |
| Age | -0.0024 [-0.0030, -0.0018] | **<0.001** | 872 | -0.0027 [-0.0034, -0.0020] | **<0.001** | 712 |
| Anxiety*Age | -0.0001 [-0.0002, -2.1e-05] | **0.015** | 881 | -0.0001 [-0.0002, -5.9-06] | **0.038** | 691 |
| Self-esteem | -0.0014 [-0.0018, -0.0010] | **<0.001** | 1245 | -0.0012 [-0.0016, -0.0007] | **<0.001** | 960 |
| Age | -0.0024 [-0.0030, -0.0018] | **<0.001** | 884 | -0.0027 [-0.0034, -0.0020] | **<0.001** | 716 |
| Self-esteem*Age | 0.0001 [2.9e-05, 0.0002] | **0.011** | 890 | 0.0001 [9.6e-06, 0.0002] | **0.034** | 690 |
| Psychological resilience[b] | | | | | | |
| Resilient | -0.0085 [-0.0157, -0.0014] | **0.019** | 1128 | -0.0051 [-0.0128, 0.0026] | 0.20 | 922 |
| Wellbeing without adversity | -0.0079[-0.0165, 0.0007] | 0.07 | 1138 | -0.0060 [-0.0152, 0.0026] | 0.21 | 902 |
| Distress without adversity | -0.0101 [-0.0259, 0.0057] | 0.21 | 1115 | -0.0128 [-0.0295, 0.0039] | 0.14 | 903 |
| Age | -0.0044 [-0.0060, -0.0029] | **<0.001** | 832 | -0.0052 [-0.0070, -0.0034] | **<0.001** | 694 |
| Resilient*Age | 0.0020 [0.0002, 0.0038] | **0.027** | 828 | 0.0027 [0.0007, 0.0046] | **0.009** | 686 |
| Wellbeing*Age | 0.0032 [0.0011, 0.0052] | **0.003** | 812 | 0.0035 [0.0012, 0.0058] | **0.003** | 671 |
| Distress*Age | 0.0006 [-0.0033, 0.0044] | 0.77 | 812 | 0.0028 [-0.0014, 0.0070] | 0.20 | 676 |

Model 1 adjusted for sex and age. Model 2 adjusted for sex, age, puberty status at baseline, physical activity, dietary pattern at baseline, caregivers' education level at baseline, and caregiver's BMI.

*Indicates interaction term.

[a] Depressive symptoms, anxiety symptoms, self-esteem and age were centered around their respective sample means before running the linear mixed models.

[b] Reference = non-resilient group. Coefficients for other groups represent the differences relative to this group.

**Supplementary Table 5**. Results from pairwise comparisons of groups according to levels of depression and anxiety symptoms, self-esteem and psychological resilience on change between baseline and follow-up in WtHr (Δ WtHr).

| Mental health indicator | Comparison | Model 1: ΔWtHr | *p*-value | Model 2: ΔWtHr | *p*-value |
|---|---|---|---|---|---|
| Depressive symptoms | | | | | |
| | Low vs. Average | 0.0011 | 0.24 | 0.0017 | 0.13 |
| | High vs. Average | -0.0029 | **0.001** | -0.0028 | **0.004** |
| | High vs. Low | -0.0041 | **0.001** | -0.0045 | **0.001** |
| Anxiety symptoms | | | | | |
| | Low vs. Average | 0.0002 | 0.87 | 0.0002 | 0.84 |
| | High vs. Average | -0.0011 | 0.19 | -0.0017 | 0.08 |
| | High vs. Low | -0.0013 | 0.27 | -0.0019 | 0.15 |
| Self-esteem | | | | | |
| | Low vs. Average | -0.0016 | 0.08 | -0.0027 | **0.010** |
| | High vs. Average | 0.0012 | 0.16 | 0.0008 | 0.36 |
| | High vs. Low | 0.0027 | **0.012** | 0.0035 | **0.005** |
| Psychological resilience | | | | | |
| | Resilient vs. Non-resilient | 0.0020 | **0.027** | 0.0027 | **0.009** |
| | Resilient vs. Wellbeing | -0.0012 | 0.14 | -0.0008 | 0.35 |
| | Resilient vs. Distress | 0.0014 | 0.43 | -0.0001 | 0.96 |
| | Non-resilient vs. Wellbeing | -0.0032 | **0.003** | -0.0035 | **0.004** |
| | Non-resilient vs. Distress | -0.0006 | 0.77 | -0.0028 | 0.20 |
| | Wellbeing vs. Distress | 0.0026 | 0.17 | 0.0007 | 0.73 |

Δ = Difference in b coefficient.

Model 1 adjusted for sex and age. Model 2 adjusted for sex, age, puberty status at baseline, physical activity, dietary pattern at baseline, caregivers’ education level at baseline, and caregiver’s BMI.

Low group = mean - 1 SD, High group = mean + 1 SD.

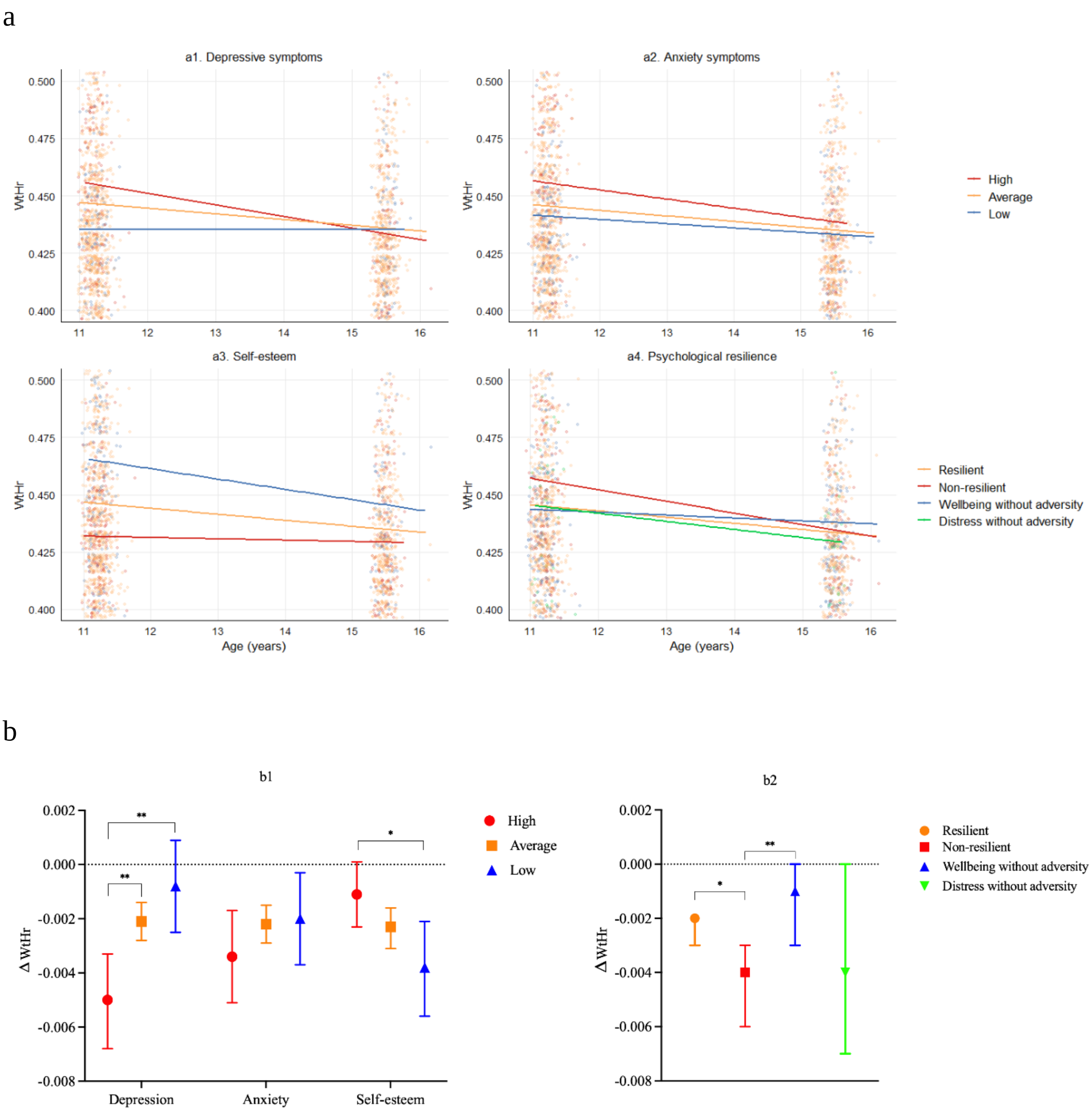


**Supplementary figure 1.** Panels A1–A4 illustrate the interaction between age x (A1) depressive symptoms, (A2) anxiety symptoms, (A3) self-esteem, or (A4) psychological resilience predicting changes in WtHr, corresponding to the terms reported in Supplementary Table 3.

Panels B1 and B2 present yearly changes in WtHr with 95% confidence intervals, alongside pairwise group comparisons: low, average and high levels of depressive and anxiety symptoms, self-esteem (B1) and different resilience groups (B2).

For depressive symptoms, anxiety symptoms, and self-esteem, participants were grouped into low (mean - 1 SD), average, and high (mean +1 SD).

Individual raw data points are overlaid in panels A1-A4 to illustrate distribution of observations.

The y-axis range is restricted to 0.40-0.50 in panel A and -0.008-0.002 in panel B to better illustrate the modest differences over time. Positive and negative beta values indicate the direction of associations.

Pairwise comparisons between the low, average, and high groups were conducted using the *emmeans* package in R (see Supplementary Table 6). *$p<0.05$, **$p<0.01$.
